# Fast and frugal decision tree for the rapid critical appraisal of systematic reviews

**DOI:** 10.1101/2023.03.20.23287481

**Authors:** Robert C. Lorenz, Mirjam Jenny, Anja Jacobs, Katja Matthias

## Abstract

Conducting high-quality overviews of reviews (OoR) is time-consuming. Because the quality of systematic reviews (SRs) varies, it is necessary to critically appraisal SRs when conducting an OoR. A well-established appraisal tool is AMSTAR 2, which takes about 15 to 32 minutes per application. To save time, we developed two fast-and-frugal decision trees (FFTs) for assessing the methodological quality of SR for OoR either during the full text screening stage (Screening FFT) or to the resulting pool of SRs (Rapid Appraisal FFT).

To build a data set for developing the FFT, we identified published AMSTAR 2 appraisals. Overall confidence ratings of the AMSTAR 2 were used as criterion and the 16 items as cues.

1,519 appraisals were obtained from 24 publications and divided into training and test data sets. The resulting Screening FFT consists of three items and correctly identifies all non-critically low-quality SRs (sensitivity of 100%), but a positive predictive value of 59%. The three-item Rapid Appraisal FFT correctly identifies 80% of the high-quality SRs and correctly identifies 97% of the low-quality SRs resulting in an accuracy of 95%. When applying the FFTs about 10% of the 16 AMSTAR 2 items are used.

The Screening FFT may be applied during full text screening in order to exclude SRs with critically low quality. The Rapid Appraisal FFT may be applied to the final SR pool to identify SR that might be of high methodological quality.

## Introduction

Policy makers in health care systems often have to make decisions based on the one hand on broad and high-quality evidence and on the other hand within limited time periods. Ideally, systematic reviews (SRs) summarize the best available evidence and serve as a basis for an informed decision. An overview of reviews (OoR) can help to identify the most relevant SRs for a specific research question. Such overviews are also called umbrella reviews, metareviews or systematic reviews of reviews. Along with the steadily increasing number of SRs (Bastian et al., 2010; Hoffmann et al., 2021; Niforatos et al., 2019), the number of overviews of reviews is also increasing (Bougioukas et al., 2021; De Santis et al., 2021; Lunny et al., 2022). Even the recent version of the Cochrane Handbook includes a chapter about overviews of reviews (Pollock et al., 2021) showing that the conduct of such an OoR should be done very carefully. Due to varying quality of SRs (Ioannidis, 2016), their critical appraisal is necessary and recommended by the Cochrane Handbook (Pollock et al., 2021). Moreover, assessing the methodological quality of SRs in OoR is also defined in an item in the recently published PRIOR statement, a reporting guideline for OoR of healthcare interventions (Gates et al., 2022). There are already many tools available for the assessment of the methodological quality of SRs (Editors, 2015; Zeng et al., 2015), although there is no consensus on the most suitable appraisal instrument for conducting an OoR (Gates et al., 2020). One of the commonly applied critical appraisal tools in earlier overviews was ‘A Measurement Tool to Assess Systematic Reviews’ (AMSTAR) (Shea et al., 2009), which was updated in 2017 (AMSTAR 2, Shea et al., 2017) and is applied in 2020 publications more often than the first version of AMSTAR (Bojcic et al., 2022). The valid and moderately reliable AMSTAR 2 (Gates et al., 2020; Lorenz et al., 2019; Pieper et al., 2019) comprises 16 items and a resulting overall confidence rating. When there are many SRs available that answer a specific research question, it is time consuming to critically appraise each SR with 16 items. The completion time for the appraisal for one SR with AMSTAR 2 ranged from 15 to 32 minutes in the original publication (Shea et al., 2017). Comparable time requirements (mean times ranged between 18 to 24 minutes) were also reported by other authors (Dang et al., 2020; Pieper et al., 2019).

In the current study, we aim to shorten the time needed to critically appraise the methodological quality of SRs using AMSTAR 2, ideally without compromising accuracy. One way to speed up critical appraisal is to use a combination of the best predictive items for the judgement in the overall confidence ratings. To this end, frugal decision trees (FFTs) can serve as powerful tools in terms of accuracy and speed with the additional benefits that they are structured simply and easy to use (Martignon et al., 2008; Martignon et al., 2003).

FFTs could simplify the critical appraisal of SRs at two different stages of conducting an OoR. It can be applied at an early stage, namely during the screening process in order to exclude SRs with very low methodological quality or it can be applied to the resulting pool of SRs in order to highlight the SRs with the highest methodological quality. Thus, the goal of the current study is to create two FFTs, one for the screening process and another for the rapid appraisal of methodological quality.

## Materials and Methods

### Original AMSTAR 2 tool for the critical appraisal of the methodological quality

The aim of the current study is to create a decision-tree based version of the AMSTAR 2 (Shea et al., 2017). This critical appraisal tool consists of 16 items and the overall confidence rating. Items are evaluated either with ‘‘Yes’’ or ‘‘No’’ (items 1, 3, 5, 6, 10, 13, 14, and 16); with ‘‘Yes’’, ‘‘Partial Yes’’, or ‘‘No’’ (items 2, 4, 7, 8, and 9); or with ‘‘Yes’’, ‘‘No’’, or ‘‘No meta-analysis conducted’’ (items 11, 12, and 15). For the overall confidence rating, the response options are ‘‘High’’, ‘‘Moderate’’, ‘‘Low,’’ and ‘‘Critically low’’.

According to the original publication (Shea et al., 2017), the items 2 (pre-defined protocol), 4 (search strategy), 7 (list of excluded studies), 9 (assessment of risk of bias), 11 (appropriate meta-analysis methods), 13 (adequate discussion of risk of bias), and 15 (publication bias) serve as critical items (seven items in total) for the overall confidence rating. A negative appraisal for a critical item is a “critical flaw”, whereas a negative appraisal for a non-critical item is a “non-critical weakness”. According to box 2 of the AMSTAR 2 publication (Shea et al., 2017) one critical flaw leads to a “low” quality rating and more than one critical flaw to a “critically low” quality rating. For a “high” quality appraisal one non-critical weakness is allowed, whereas more than one non-critical weakness leads to a “moderate” quality appraisal.

### Study design

A fast and frugal decision tree can yield a fast and accurate decision in a complex situation based on a limited amount of information (Martignon et al., 2008; Martignon et al., 2003). An FFT is constructed with few highly predictive cues (here AMSTAR 2 items) and leads to a binary decision (here a dichotomized AMSTAR 2 overall confidence rating). The FFT has one exit leaf on every level of the tree (here an AMSTAR 2 item), i.e. for every item, one of its outcomes can lead to a decision (Phillips et al., 2017). In other words: with an FFT the response to a single AMSTAR 2 item might be enough to classify a SR into high or low methodological quality.

Constructing a robust data-induced FFT requires training and test data. In the current study, already available data mostly from published OoR applying the AMSTAR 2 is used. In order to collect the data, a systematic literature search was conducted (see details below). AMSTAR 2 data was extracted, prepared, and split into a training and test data set to train and test the FFT. We selected the resulting FFTs to meet the requirements of two different goals: An FFT for screening purposes and another FFT for a rapid appraisal of methodological quality.

### Literature search and study selection

Electronic searches for bibliographic records containing the term AMSTAR 2 or variants thereof were run in: PubMed, Epistemonikos, and CINAHL via EBSCO in September 2020. The search strategy including key words is described in the supplemental material (see Table S1). All records were imported into an electronic database (EndNote, X9) and deduplicated.

Studies were selected a two-stage screening process. The initial screening of titles and abstracts was conducted using the inclusion and exclusion of criteria as defined below. For the resulting set of studies, study reports were obtained in full-text and checked in detail for eligibility. Each stage of the screening process was conducted independently by the authors KM and RL. Each case of disagreement was discussed until consensus was reached.

Studies were eligible when (1) the AMSTAR 2 appraisal tool was applied to a minimum of 20 SRs. Further, (2) for the overall confidence rating of the AMSTAR 2, the appraisal scheme as suggested by the original AMSTAR 2 article (Shea et al., 2017) had to be applied (see details in the AMSTAR 2 description), i.e. the the application of the AMSTAR 2 website scheme did not qualify because this appraisal scheme seems to lead to different results (Lorenz et al., 2021; Pieper et al., 2021). Studies that applied another appraisal scheme or did not report an overall confidence rating were excluded. (3) If the data of the AMSTAR appraisal for each SR was not available in the fulltext (including supplemental material), the authors of the study were contacted. In case of no response, a reminder was sent. When the data was still not availble, the study was excluded. (4) Retrieved AMSTAR 2 appraisals were checked for plausibility. All SRs with an overall confidence rating of “moderate” or “high” quality were checked. According to the original publication (Shea et al., 2017) one negative evaluation (we used a “no” response) leads to a low or critically low quality rating. If this rule was violated in more than 10 percent of the moderate and high-quality ratings, the publication was excluded. See supplementary Figure S1 for a study flow diagram.

### Data extraction

Most of the included publications contained tables with the AMSTAR 2 appraisals for each SR. In those cases, tables were transformed automatically into Microsoft Excel format by using Adobe Acrobat Pro DC software. Excel sheets were then formatted into a template containing responses to the 16 AMSTAR 2 items and the overall confidence rating. This procedure was applied in order to avoid manual transmission errors.

### Data analysis

The FFTs were developed with the fast-and-frugal decision trees toolbox FFTrees (Phillips et al., 2017) in the software environment and programming language R (R Core Team). The overall confidence rating had to be dichotomized. As we aimed to develop two different FFTs, dichotomization was done differently for each purpose (screening and appraisal). For the Screening FFT “critically low” quality ratings were assigned to the very low category and the three other quality ratings (“high”, “moderate”, and “low”) were assigned to non-critically low category. For the Rapid Appraisal FFT, the four categories were dichotomized into high quality (“high” and “moderate” AMSTAR 2 appraisal) and low quality (“low” and “critically low” AMSTAR 2 appraisal).

The inter-rater reliability of AMSTAR 2 is far from perfect and mostly described as moderately reliable (Gates et al., 2020; Lorenz et al., 2019; Pieper et al., 2019). We assume that different AMSTAR 2 users apply AMSTAR 2 slightly differently. In order to keep the varying appraisal strategies consistent across the training and test set, we balanced the distribution of overall confidence ratings. To this end, we stratified AMSTAR 2 appraisals of each SR according to the dichotomized criterion for each tree. This procedure is concretely described in the following example: in case of the Rapid Appraisal FFT, we randomly drew from an OoR with 30 individual critical appraisals containing 4 SRs with “high” quality and 26 SRs with “low” quality, and assigned 2 SRs with “high” quality and 13 SRs with “low” quality to the test and training data sets, respectively. We repeated this procedure for each of the included OoR and concatenated all resulting AMSTAR 2 appraisals in well-balanced test and training data sets for the development of the two trees (Screening FFT and Rapid Appraisal FFT).

The FFTrees toolbox produces many FFTs with different characteristics. The accuracy for each tree can be evaluated based on a two-by-two cross table summarizing the correct and incorrect decisions of the tree split for high and low-quality SRs. In the terminology of signal detection theory, this table contains hits, false alarms, misses, and correct rejections. In the case of the Rapid Appraisal FFT, a hit is a correctly identified high-quality SR, a false alarm has a high-quality SR according to FFT, but truly has low quality, a correct rejection is a correctly identified low-quality SR, and a miss is a low-quality SR according to FFT, but truly has high quality. In case of the Screening FFT, a hit is a correctly identified non-critically low-quality SR, a false alarm is a non-critically low-quality SR according to FFT, but truly has critically low quality, a correct rejection is a correctly identified critically low-quality SR, and a miss is a critically low-quality SR according to FFT, but truly has non-critically low quality.

Different measures of performance can be calculated based on the table: the FFT’s sensitivity (proportion of correctly identified true positive cases or hits), specificity (proportion of correctly identified true negative cases or correct rejections), positive predictive value (proportion of all SRs which the FFT deems high/non-critically quality and who are truly high/non-critically quality), and negative predictive value (proportion of all SRs which the FFT deems low/critically low quality and which are truly low/critically low quality). Furthermore, the overall accuracy is defined as the overall percentage of correct decisions (number of hits and correct rejections divided by number of all decisions). This accuracy definition ignores the fact that the hits might considerably outnumber the correct rejections or vice versa. To take potential differences in the number of hits and correct rejections into account, balanced accuracy is defined as the average of the sensitivity and the specificity.

The FFTrees toolbox also estimates decision speed and frugality. Mean cues used (mcu) computes the average number of cues (number of AMSTAR 2 items) used for decision making averaged across all cases (SRs). Another indicator of a tree’s frugality is the percentage of cues ignored (pci), which is defined as 1 minus mcu divided by the total number of cues in the dataset (in this case 16). The pci value, therefore, represents a quantitative metric for the ignored information, i.e. unused AMSTAR 2 items.

The Screening FFT should avoid misses, i.e., SRs that are classified as critically low quality by the FFT, but according to AMSTAR2 appraisal are classified as non-critically low quality. Since the number of misses is considered in the sensitivity measure, sensitivity was used to identify the best Screening FFT. For the Rapid Appraisal FFT, an FFT with the best overall performance, i.e. the highest accuracy, is the ideal FFT. Thus, we used that metric to select the FFT. Additionally, the FFT with the highest balanced accuracy is presented in the supplemental material. Please note that both FFTs, the Screening FFT and the Rapid Appraisal FFT, were both developed and tested on the whole data set, but independently from each other.

Apart from FFT analyses we calculated descriptive statistics of the AMSTAR 2 ratings across all included critical appraisals of the SRs.

## Results

### Study pool

Our search retrieved 270 references after deduplication. Of these 71 study reports were obtained and screened in full-text for eligibility. In 20 cases the original overall confidence rating as proposed by Shea et al. (2017) was not applied, in another 7 cases no overall rating was reported (in most of these cases the authors preferred a sum score across the AMSTAR 2 items). In another 7 cases, data was not available even upon repeated request to the authors. The resulting study pool contained 24 publications with 1,519 AMSTAR 2 appraisals. Included studies encompassed various fields including oncology, urology, telemedicine, psychology, and orthopaedics. The study flow diagram (Figure S1) as well as the full list of included publications can be found in the supplementary material.

### Screening FFT

The purpose of the Screening FFT was to exclude SRs with “critically low” methodological quality. We selected the FFT with the highest sensitivity. This tree (see Figure 1) starts with the cue that checks for a pre-defined protocol (item 2, exit the tree and decide “non-critically low” at a partial yes or yes). The next cue checks for a list of excluded studies (item 7, exit the tree and decide “critically low” at a no). The final cue concerns the discussion of risk of bias in the results of the review (item 13, decide “critically low” at a no and “non-critically low” at a yes).

**Figure 1:**
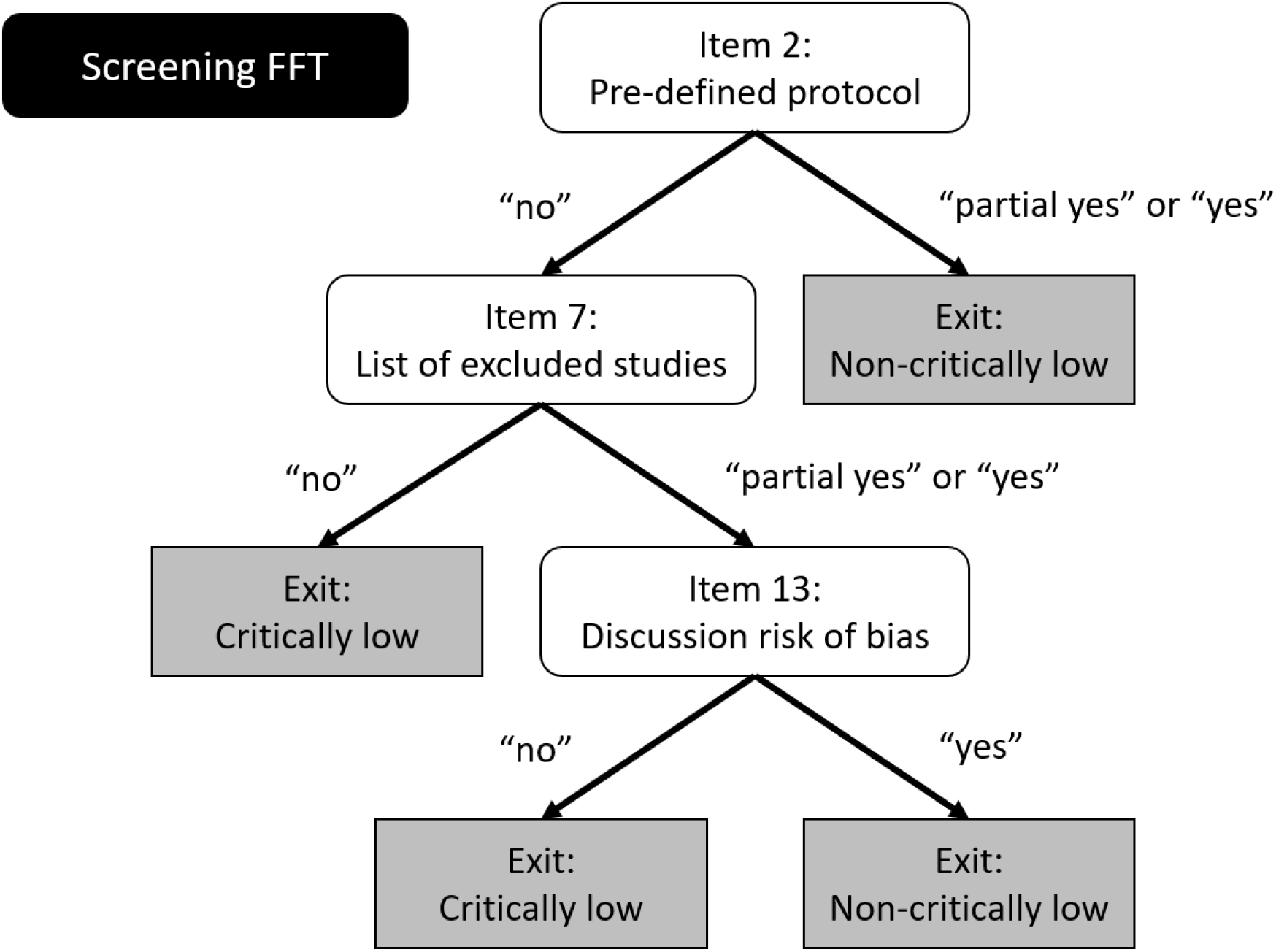
The fast-and-frugal decision tree for screening purposes. This tree contains three items of the AMSTAR 2 that are structured in the following way: On the first level, item 2 (pre-defined protocol), on second level item 7 (list of excluded studies) and on final level item 13 (discussion of risk of bias).

According to AMSTAR 2 appraisals 25 % of the SRs were of non-critically low quality. When applied to the test data set, which consisted of 762 cases, the Screening FFT produced 0 misses and 193 hits and, therefore, had a sensitivity of 100%. It had a specificity of 77% with 436 correct rejections and 133 false alarms. The positive predictive value was 59% indicating that a classification as high quality based on the FFT was correct in about 6 out of 10 cases. The negative predictive value was 100%, suggesting that a classification as low quality based on the FFT was always correct. The overall accuracy was 83% and the balanced accuracy was 88%.

The measurements of speed and frugality were 1.7 mcu, i.e. on average two items were necessary to decide. The pci was 90%, i.e. on average 90% of the AMSTAR 2 items were ignored.

### Rapid appraisal FFT

To rapidly identify SR of potentially high quality, the FFT with the highest accuracy was selected to maximize hits and correct rejections. This decision tree (see Figure 2) also contains AMSTAR 2 items 2 and 7 (as does the Screening FFT), but starts with item 7 about the list of excluded studies (exit the tree and decide “low” at a no), that is followed by item 2 about the protocol (exit the tree and decide “low” at a no or partial yes). Finally, the last leave of the tree contains item 4 that checks for a search strategy (exit the tree and decide “low” at a no or partial yes and exit “high” at a yes).

**Figure 2:**
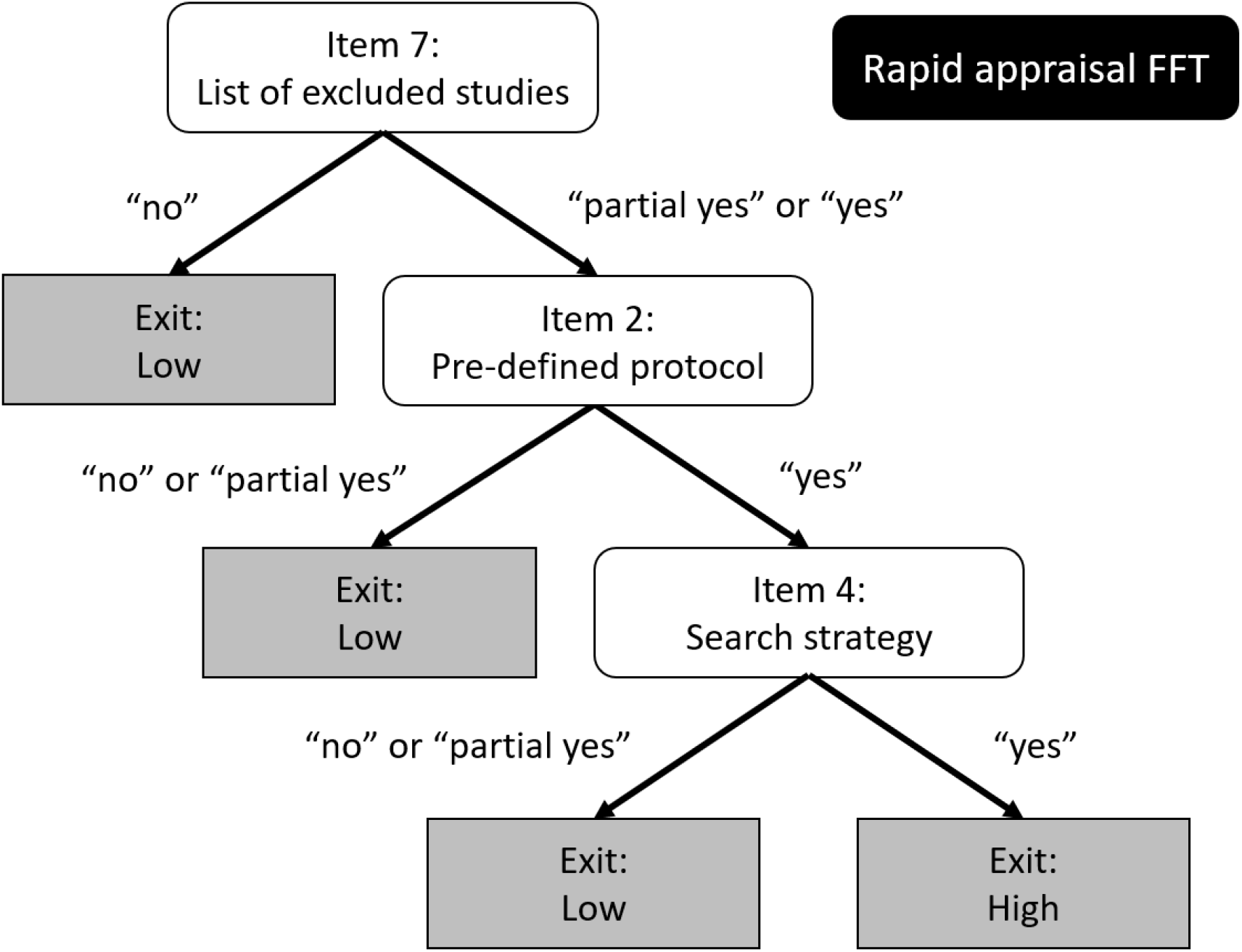
The fast-and-frugal decision tree for short appraisal. This tree contains three items of the AMSTAR 2 that are ordered the following way: On the first level item 7 (list of excluded studies), on second level item 2 (pre-defined protocol) and on final level item 4 (search strategy).

Only 12 % of the included AMSTAR 2 appraisals resulted in high quality (comprised “high” and “moderate” AMSTAR 2 appraisals), whereas 88% resulted in low quality (comprised “low” and “critically low” AMSTAR 2 appraisals). The FFT’s accuracy in the test data set (which contained 758 cases) was 95% with 75 hits, 647 correct rejections, 19 misses and 17 false alarms leading to a sensitivity of 80% and a specificity of 97%. The positive predictive value was 82% and the negative predictive value was 97%. In this FFT the mcu was 1.4, i.e. on average, 1 item was necessary to decide and the pci was 91%, i.e. on average 91% of the AMSTAR 2 items were ignored.

Since the sensitivity in this case was 80%, about 2 out of 10 SR were falsely classified as low quality, we also attached a decision tree in the supplementary material with the highest balanced accuracy (92%) that weights sensitivity and specificity equally. This alternative FFT had a sensitivity of 95% and a specificity of 91% (5 misses), which was at the expense of a positive predictive value of only 60%

### Summary descriptive statistics of AMSTAR 2 appraisals

The descriptive statistics were aggregated across all 24 included publications (see Figure 3). The items that are part of the decision trees are particularly important. The protocol item (item 2) was rated with yes in 27%, partial yes in 10%, and with no in 63%. The item about the list of excluded studies (item 7) was rated with yes in 25%, partial yes in 4%, and with no in 71%. The item about the discussion of risk of bias (item 13) of the Screening FFT was rated with yes in 56% and with no in 44%. The item about the search strategy (item 4) of the Rapid Appraisal FFT was rated with yes in 26%, partial yes in 50%, and with no in 24%.

**Figure 3:**
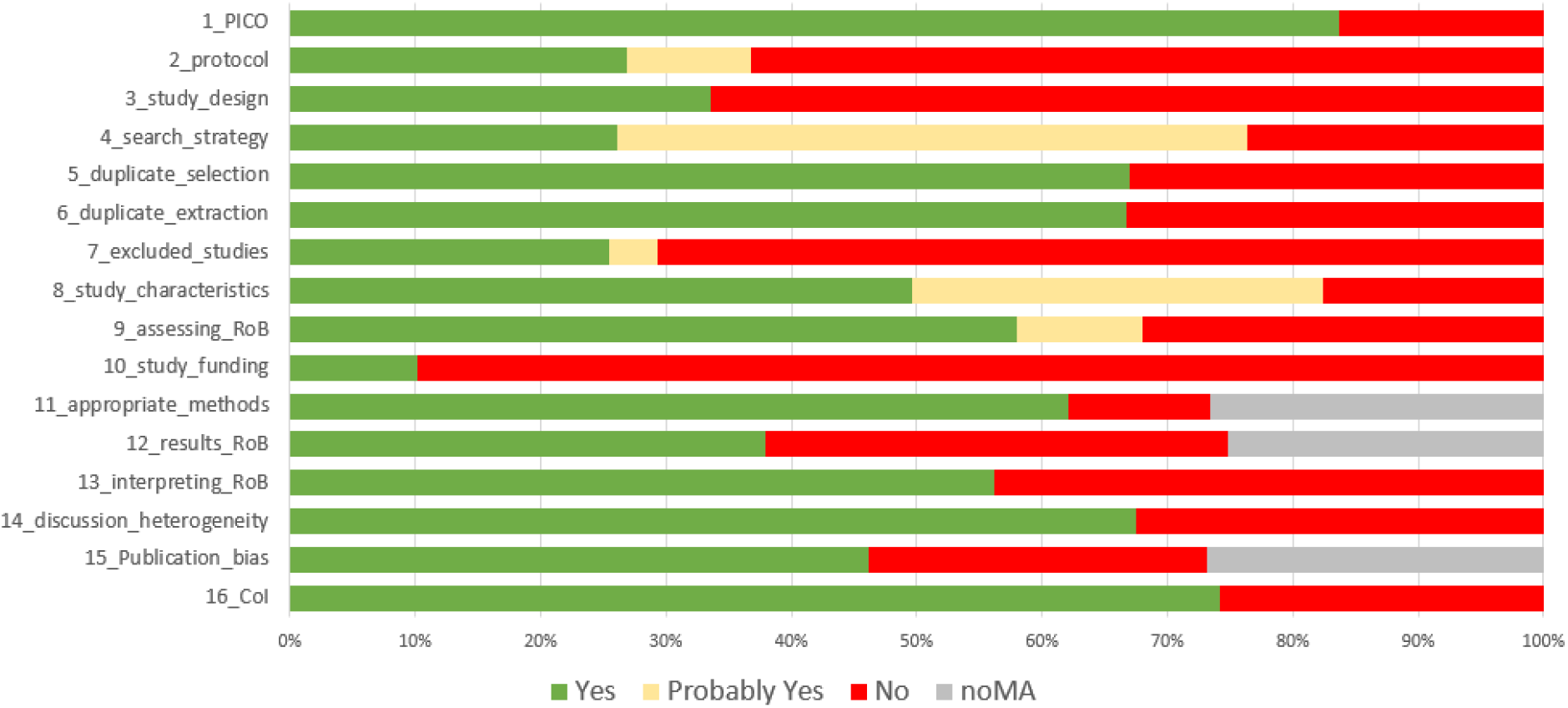
Summary descriptive statistics of AMSTAR 2 appraisals for each of the 16 items of the AMSTAR 2 across all 24 studies including together 1519 critical appraisals.

## Discussion

The aim of the current study was to develop two fast-and-frugal decision trees (FFTs) to rapidly assess the methodological quality that could be applied to the preparation of an OoR either during the full text screening stage (Screening FFT) or to the resulting pool of SRs (Rapid Appraisal FFT). The FFTs were trained and tested on a dataset that contains 1,519 AMSTAR 2 critical appraisals that were obtained from 24 publications. The Screening FFT contains three items, has a sensitivity of 100%, and a positive predictive value of 59% (overall accuracy: 83%). The Rapid Appraisal FFT also contains three items, has a sensitivity of 80% and a specificity of 97% resulting in an overall accuracy of 95%. When applying the FFTs, about 10% of the 16 AMSTAR 2 items are used, i.e. the vast majority is ignored.

### Applying the Screening FFT

The Screening FFT was designed to identify “critically low” quality SRs. In the test data set no misses were observed leading to a high sensitivity of 100%. However, when the Screening FFT deemed an SR “non-critically low”, this appraisal was correct in only 6 out of 10 cases. In practice, we suggest to implement the Screening FFT during the full text screening process, e.g. as an eligibility criterion. When the result of the Screening FFT is “non-critically low”, we recommend to apply the full AMSTAR 2 checklist with all 16 items (or at least the seven critical items) and the overall confidence rating in order to get valid appraisal. This suggested step-by-step application of the AMSTAR 2 may save a lot of time during the preparation of an OoR.

### Applying the Rapid Appraisal FFT

The Rapid Appraisal FFT has a high accuracy of 95%. The potential miss-classification of 5% of the reviews is detrimental if these contain the most relevant evidence. Therefore, the Rapid Appraisal FFT gives only a tentative hint whether an SR is of high or low methodological quality. Importantly, by applying this tree, more than 90% of the AMSTAR 2 ratings could be ignored indicating a large amount of potentially saved time. The Rapid Appraisal FFT may therefore be used in situations where the time pressure is very high and the number of SRs is enormous.

### Predictive value of items of the FFTs

Both decision trees contain the items 2 (pre-defined protocol) and 7 (excluded studies), which were both critical items for the overall confidence rating of the AMSTAR 2 as suggested by Shea et al. (Shea et al., 2017). According to item 2, in the current sample, nearly two thirds of the SRs were rated with a “no” indicating that no pre-specified protocol is available. Previous studies have shown that SRs with a published protocol were more elaborated, reported more thoroughly (Allers et al., 2018), and have a higher methodological quality (Ge et al., 2018; Sideri et al., 2018). Additionally, Cochrane reviews usually publish a protocol a priori. Thus, the item has a high predictive value for the overall methodological quality of a SR.

Item 7 assesses whether a list of excluded studies is provided in the SR (ideally justification for each SR). The majority of the included SRs (71%) in this study did not provide such a list. A recent study identified item 7 as one of the main critical items and therefore triggers for a low or critically low overall rating in the AMSTAR 2 (Siemens et al., 2021). Similar to item 2, Cochrane reviews usually provide a list of excluded studies including a justification of each exclusion.

The Screening FFT also includes the critical item 13 (Shea et al., 2017) which requires an adequate interpretation or discussion of the risk of bias of the individual studies. In the current sample more than half of the SRs fulfilled this item (56%). In contrast to item 2 and 7, this item 13 is not related to the methodical conduction of a SRs and a kind of reporting quality. To the best of our knowledge there are no studies that investigated the predictive value of this item 13 on the overall methodological quality of SRs.

Finally, the Rapid Appraisal FFT includes the critical item 4 that assesses whether the authors of the SR applied a comprehensive literature search strategy. This item was rated with yes in 26%, partial yes in 50%, and with no in 24% of cases in the current sample. The search strategy is one of the key issues when conducting a systematic review. A PRISMA extension for reporting of literature searches in SRs was recently published (PRISMA-S, Rethlefsen et al., 2021). Previous studies showed that involving librarians or information specialists may improve the reproducibility of literature searches (Koffel, 2015; Rethlefsen et al., 2015) and may be associated with a lower risk of bias of the SRs as shown in a small sample (Aamodt et al., 2019). Another study showed that including information about the search for unpublished studies is related to the methodological quality of SRs (Storman et al., 2022). The fact that the search strategy item is also included in the Rapid Appraisal FFT additionally highlights the role of a high-quality literature search on the overall quality appraisal of SRs.

### Methods to reduce costs in evidence synthesis

SRs and overviews of reviews are important for policy makers to make informed decisions. However, high-quality comprehensive evidence syntheses cost a significant amount of time and resources (Clark et al., 2020). It is often not feasible to conduct such SRs in short time frames (Lagisz et al., 2018). In rapid reviews, short-cuts or automation tools are used to save time. Time could be saved e.g. by restricting the number of databases (only two data bases) or search years (limit to the past five years) in the literature search, not performing full parallel screening (Lagisz et al., 2018), rule-based approaches for automatic data extraction (Bashir et al., 2021), reducing synthesis depth (e.g. conducting a narrative review) (O’Leary et al., 2017), or even using automation tools like RobotReviewer to automatically assess risk of bias in randomized controlled trials (Marshall et al., 2017; Marshall et al., 2016). The Screening FFT may be used to conduct OoR that include only non-critically low SRs. This is similar to a recent OoR (Smith et al., 2019) that only included non-critically low SRs based on the full AMSTAR 2 appraisal.

### Limitations

The current study has some limitations. First, the FFTs are based on AMSTAR 2 appraisals from the included publications. We included only studies that applied the suggested overall confidence rating scheme that is based on seven critical items as suggested by Shea et al. (Shea et al., 2017). However, as suggested by Shea et al. the rating scheme may also be adjusted by AMSTAR 2 users to their specific research question. Therefore, FFTs should only be applied to research questions that are also appropriate to the seven-item scheme. Second, the interrater reliability (IRR) of the AMSTAR 2 varies across the items and this variation is also present in the input data for the FFTs. Specifically, the limited IRR of some AMSTAR 2 items may negatively affect the results. The IRR of items 2 and 7 are relatively high (item 2: substantial to almost perfect, item 7: moderate to substantial), whereas the IRR of items 4 and 13 are predominantly fair (item 4: fair, item 13: fair to moderate) (Lorenz et al., 2019; Pieper et al., 2019). Third, the literature search in the current study is more than two years old. In the meantime, more overviews of reviews with AMSTAR 2 appraisals may have been published. We also included only overviews of reviews that included more than 20 SRs. Nevertheless, we were able to include more than 1,500 critical appraisals that represents already a considerable data set. Fourth, items 2 and 7 are usually fulfilled in Cochrane Reviews. Thus, both FFT may reliably separate Cochrane reviews from non-Cochrane reviews.

## Conclusions

We developed two FFTs in that save resources during the preparation of an OoR. On the one hand, the Screening FFT may be applied during full text screening in order to exclude SRs with critically low quality. On the other hand, the Rapid Appraisal FFT may be applied to the final SR pool to identify SR that might be of high methodological quality. Another possibility may be a two-staged approach, first applying the Screening FFT to exclude SRs with non-critically low quality and subsequently apply the full AMSTAR 2 to the remaining SRs. These approaches may not only be used in the conduction of an OoR, but also in the development of clinical guidelines.

## Data Availability

The data set in this study is available from the corresponding author upon reasonable request.

## Acknowledgements

We thank Lydia Jones for assisting with language editing and proofreading. We furthermore thank Dr. Konrad Neumann (Charité Berlin, Institute of Biometry and Clinical Epidemiology) for discussing about the research design. Finally, we thank all authors, who already published AMSTAR 2 ratings as well as those, who provided data upon request.

## Data availability statement

The data set in this study is available from the corresponding author upon reasonable request.

## Supplementary data

Supplementary material is available.

## Authors’ contributions

RL, AJ and KM conceived of the study and participated in its design and coordination. RL performed the analysis under the supervision of MJ. RL and KM participated in data collection and all authors discussed and interpreted the data. RL drafted the manuscript. All authors read and approved the final manuscript.

## Funding

This research was not supported by any grant from funding agencies in the public, commercial, or not-for-profit sectors.

## Conflict of Interest

The authors declare to have no conflicts of interest.

## Ethics approval statement

Ethics approval was not required for this study, because only published data was used for data analysis.

## Notes

### Competing Interest Statement

The authors have declared no competing interest.

## References

Aamodt, M., Huurdeman, H., & Strømme, H. (2019). Librarian Co-Authored Systematic Reviews are Associated with Lower Risk of Bias Compared to Systematic Reviews with Acknowledgement of Librarians or No Participation by Librarians. Evidence Based Library and Information Practice, 14, 103–127.

Allers, K., Hoffmann, F., Mathes, T., & Pieper, D. (2018). Systematic reviews with published protocols compared to those without: more effort, older search. J Clin Epidemiol, 95, 102–110. https://doi.org/10.1016/j.jclinepi.2017.12.005

Bashir, R., Dunn, A. G., & Surian, D. (2021). A rule-based approach for automatically extracting data from systematic reviews and their updates to model the risk of conclusion change. Res Synth Methods, 12(2), 216–225. https://doi.org/10.1002/jrsm.1473

Bastian, H., Glasziou, P., & Chalmers, I. (2010). Seventy-five trials and eleven systematic reviews a day: how will we ever keep up? PLoS Med, 7(9), e1000326. https://doi.org/10.1371/journal.pmed.1000326

Bojcic, R., Todoric, M., & Puljak, L. (2022). Adopting AMSTAR 2 critical appraisal tool for systematic reviews: speed of the tool uptake and barriers for its adoption. BMC Med Res Methodol, 22(1), 104. https://doi.org/10.1186/s12874-022-01592-y

Bougioukas, K. I., Vounzoulaki, E., Mantsiou, C. D., Papanastasiou, G. D., Savvides, E. D., Ntzani, E. E., & Haidich, A. B. (2021). Global mapping of overviews of systematic reviews in healthcare published between 2000 and 2020: a bibliometric analysis. J Clin Epidemiol, 137, 58–72. https://doi.org/10.1016/j.jclinepi.2021.03.019

Clark, J., Glasziou, P., Del Mar, C., Bannach-Brown, A., Stehlik, P., & Scott, A. M. (2020). A full systematic review was completed in 2 weeks using automation tools: a case study. J Clin Epidemiol, 121, 81–90. https://doi.org/10.1016/j.jclinepi.2020.01.008

Dang, A., Chidirala, S., Veeranki, P., & Vallish, B. N. (2020). A Critical Overview of Systematic Reviews of Chemotherapy for Advanced and Locally Advanced Pancreatic Cancer using both AMSTAR2 and ROBIS as Quality Assessment Tools. Rev Recent Clin Trials. https://doi.org/10.2174/1574887115666200902111510

De Santis, K. K., Lorenz, R. C., Lakeberg, M., & Matthias, K. (2021). The application of AMSTAR2 in 32 overviews of systematic reviews of interventions for mental and behavioural disorders: A cross-sectional study. Res Synth Methods. https://doi.org/10.1002/jrsm.1532

Editors, P. M. (2015). From Checklists to Tools: Lowering the Barrier to Better Research Reporting. PLoS Med, 12(11), e1001910. https://doi.org/10.1371/journal.pmed.1001910

Gates, M., Gates, A., Duarte, G., Cary, M., Becker, M., Prediger, B., Vandermeer, B., Fernandes, R. M., Pieper, D., & Hartling, L. (2020). Quality and risk of bias appraisals of systematic reviews are inconsistent across reviewers and centers. J Clin Epidemiol, 125, 9–15. https://doi.org/10.1016/j.jclinepi.2020.04.026

Gates, M., Gates, A., Pieper, D., Fernandes, R. M., Tricco, A. C., Moher, D., Brennan, S. E., Li, T., Pollock, M., Lunny, C., Sepulveda, D., McKenzie, J. E., Scott, S. D., Robinson, K. A., Matthias, K., Bougioukas, K. I., Fusar-Poli, P., Whiting, P., Moss, S. J., & Hartling, L. (2022). Reporting guideline for overviews of reviews of healthcare interventions: development of the PRIOR statement. Bmj, 378, e070849. https://doi.org/10.1136/bmj-2022-070849

Ge, L., Tian, J. H., Li, Y. N., Pan, J. X., Li, G., Wei, D., Xing, X., Pan, B., Chen, Y. L., Song, F. J., & Yang, K. H. (2018). Association between prospective registration and overall reporting and methodological quality of systematic reviews: a meta-epidemiological study. J Clin Epidemiol, 93, 45–55. https://doi.org/10.1016/j.jclinepi.2017.10.012

Hoffmann, F., Allers, K., Rombey, T., Helbach, J., Hoffmann, A., Mathes, T., & Pieper, D. (2021). Nearly 80 systematic reviews were published each day: Observational study on trends in epidemiology and reporting over the years 2000-2019. J Clin Epidemiol, 138, 1–11. https://doi.org/10.1016/j.jclinepi.2021.05.022

Ioannidis, J. P. (2016). The Mass Production of Redundant, Misleading, and Conflicted Systematic Reviews and Meta-analyses. Milbank Q, 94(3), 485–514. https://doi.org/10.1111/1468-0009.12210

Koffel, J. B. (2015). Use of recommended search strategies in systematic reviews and the impact of librarian involvement: a cross-sectional survey of recent authors. PLoS One, 10(5), e0125931. https://doi.org/10.1371/journal.pone.0125931

Lagisz, M., Samarasinghe, G., & Nakagawa, S. (2018). Rapid reviews for the built environment – Methodology and guidelines. CRCLCL.

Lorenz, R. C., Matthias, K., Pieper, D., Wegewitz, U., Morche, J., Nocon, M., Rissling, O., Schirm, J., & Jacobs, A. (2019). A psychometric study found AMSTAR 2 to be a valid and moderately reliable appraisal tool. J Clin Epidemiol, 114, 133–140. https://doi.org/10.1016/j.jclinepi.2019.05.028

Lorenz, R. C., Pieper, D., Rombey, T., Jacobs, A., Rissling, O., Freitag, S., & Matthias, K. (2021). Reply to letter to the editor by Franco et al. AMSTAR 2 overall confidence rating: A call for even more transparency. J Clin Epidemiol, 138, 241–242. https://doi.org/10.1016/j.jclinepi.2021.03.016

Lunny, C., Neelakant, T., Chen, A., Shinger, G., Stevens, A., Tasnim, S., Sadeghipouya, S., Adams, S., Zheng, Y. W., Lin, L., Yang, P. H., Dosanjh, M., Ngsee, P., Ellis, U., Shea, B. J., Reid, E. K., & Wright, J. M. (2022). Bibliometric study of ‘overviews of systematic reviews’ of health interventions: Evaluation of prevalence, citation and journal impact factor. Res Synth Methods, 13(1), 109–120. https://doi.org/10.1002/jrsm.1530

Marshall, I. J., Kuiper, J., Banner, E., & Wallace, B. C. (2017). Automating Biomedical Evidence Synthesis: RobotReviewer. Proc Conf Assoc Comput Linguist Meet, 2017, 7–12. https://doi.org/10.18653/v1/P17-4002

Marshall, I. J., Kuiper, J., & Wallace, B. C. (2016). RobotReviewer: evaluation of a system for automatically assessing bias in clinical trials. J Am Med Inform Assoc, 23(1), 193–201. https://doi.org/10.1093/jamia/ocv044

Martignon, L., Katsikopoulos, K. V., & Woike, J. K. (2008). Categorization with limited resources: A family of simple heuristics. Journal of Mathematical Psychology, 52(6), 352–361.

Martignon, L., Vitouch, O., Takezawa, M., & Forster, M. R. (2003). Naive and yet enlightened: From natural frequencies to fast and frugal decision trees. In D. Hardman & L. Macchi (Eds.), Thinking: Psychological perspective on reasoning, judgment, and decision making (pp. 189–211). John Wiley & Sons.

Niforatos, J. D., Weaver, M., & Johansen, M. E. (2019). Assessment of Publication Trends of Systematic Reviews and Randomized Clinical Trials, 1995 to 2017. JAMA Intern Med, 179(11), 1593–1594. https://doi.org/10.1001/jamainternmed.2019.3013

O’Leary, D. F., Casey, M., O’Connor, L., Stokes, D., Fealy, G. M., O’Brien, D., Smith, R., McNamara, M. S., & Egan, C. (2017). Using rapid reviews: an example from a study conducted to inform policy-making. J Adv Nurs, 73(3), 742–752. https://doi.org/10.1111/jan.13231

Phillips, N. D., Neth, H., Woike, J. K., & Gaissmaier, W. (2017). FFTrees: A toolbox to create, visualize, and evaluate fast-and-frugal decision trees. Judgment and Decision making, 12(4), 344–368.

Pieper, D., Lorenz, R. C., Rombey, T., Jacobs, A., Rissling, O., Freitag, S., & Matthias, K. (2021). Authors should clearly report how they derived the overall rating when applying AMSTAR 2-a cross-sectional study. J Clin Epidemiol, 129, 97–103. https://doi.org/10.1016/j.jclinepi.2020.09.046

Pieper, D., Puljak, L., Gonzalez-Lorenzo, M., & Minozzi, S. (2019). Minor differences were found between AMSTAR 2 and ROBIS in the assessment of systematic reviews including both randomized and nonrandomized studies. J Clin Epidemiol, 108, 26–33. https://doi.org/10.1016/j.jclinepi.2018.12.004

Pollock, M., Fernandes, R., Becker, L., Pieper, D., & Hartling, L. (2021). Chapter V: Overviews of Reviews. In J. Higgins, J. Thomas, J. Chandler, M. Cumpston, T. Li, M. J. Page, & V. A. Welch (Eds.), Cochrane Handbook for Systematic Reviews of Interventions (Vol. 6.2). https://training.cochrane.org/handbook/current/chapter-v

Rethlefsen, M. L., Farrell, A. M., Osterhaus Trzasko, L. C., & Brigham, T. J. (2015). Librarian co-authors correlated with higher quality reported search strategies in general internal medicine systematic reviews. J Clin Epidemiol, 68(6), 617–626. https://doi.org/10.1016/j.jclinepi.2014.11.025

Rethlefsen, M. L., Kirtley, S., Waffenschmidt, S., Ayala, A. P., Moher, D., Page, M. J., Koffel, J. B., & Group, P.-S. (2021). PRISMA-S: an extension to the PRISMA Statement for Reporting Literature Searches in Systematic Reviews. Syst Rev, 10(1), 39. https://doi.org/10.1186/s13643-020-01542-z

Shea, B. J., Hamel, C., Wells, G. A., Bouter, L. M., Kristjansson, E., Grimshaw, J., Henry, D. A., & Boers, M. (2009). AMSTAR is a reliable and valid measurement tool to assess the methodological quality of systematic reviews. J Clin Epidemiol, 62(10), 1013–1020. https://doi.org/10.1016/j.jclinepi.2008.10.009

Shea, B. J., Reeves, B. C., Wells, G., Thuku, M., Hamel, C., Moran, J., Moher, D., Tugwell, P., Welch, V., Kristjansson, E., & Henry, D. A. (2017). AMSTAR 2: a critical appraisal tool for systematic reviews that include randomised or non-randomised studies of healthcare interventions, or both. Bmj, 358, j4008. https://doi.org/10.1136/bmj.j4008

Sideri, S., Papageorgiou, S. N., & Eliades, T. (2018). Registration in the international prospective register of systematic reviews (PROSPERO) of systematic review protocols was associated with increased review quality. J Clin Epidemiol, 100, 103–110. https://doi.org/10.1016/j.jclinepi.2018.01.003

Siemens, W., Schwarzer, G., Rohe, M. S., Buroh, S., Meerpohl, J. J., & Becker, G. (2021). Methodological quality was critically low in 9/10 systematic reviews in advanced cancer patients-A methodological study. J Clin Epidemiol, 136, 84–95. https://doi.org/10.1016/j.jclinepi.2021.03.010

Smith, V., Gallagher, L., Carroll, M., Hannon, K., & Begley, C. (2019). Antenatal and intrapartum interventions for reducing caesarean section, promoting vaginal birth, and reducing fear of childbirth: An overview of systematic reviews. PLoS One, 14(10), e0224313. https://doi.org/10.1371/journal.pone.0224313

Storman, D., Koperny, M., Zajac, J., Polak, M., Weglarz, P., Bochenek-Cibor, J., Swierz, M. J., Staskiewicz, W., Gorecka, M., Skuza, A., Wach, A. A., Kaluzinska, K., & Bala, M. M. (2022). Predictors of Higher Quality of Systematic Reviews Addressing Nutrition and Cancer Prevention. Int J Environ Res Public Health, 19(1). https://doi.org/10.3390/ijerph19010506

Zeng, X., Zhang, Y., Kwong, J. S., Zhang, C., Li, S., Sun, F., Niu, Y., & Du, L. (2015). The methodological quality assessment tools for preclinical and clinical studies, systematic review and meta-analysis, and clinical practice guideline: a systematic review. J Evid Based Med, 8(1), 2–10. https://doi.org/10.1111/jebm.12141

