## Supplementary Material for "Fast and frugal decision tree for the rapid critical appraisal of systematic reviews"

### *Systematic literature search*

Electronic searches took place in September 2020 in the bibliographic databases PubMed, Epistemonikos, and CINAHL via EBSCO. We used key words as shown in table S1 to identify records that contain the term “AMSTAR 2” or variants thereof. We did not apply any other restrictions.

*Table S1: search strategies*

| | |
|---|---|
| **PubMed** | amstar 2[tiab] OR amstar-2[tiab] OR amstar2[tiab] |
| **Epistemonikos** | (title(“AMSTAR 2”) OR abstract:(“AMSTAR 2”)) OR (title(“AMSTAR-2”) OR abstract:(“AMSTAR-2”)) OR (title(“AMSTAR2”) OR abstract:(“AMSTAR2”)) |
| **CINAHL via EBSCO** | TI (“amstar 2” OR amstar-2 OR amstar2) OR AB (“amstar 2” OR amstar-2 OR amstar2) |

#### ***Modified PRISMA flow diagram***

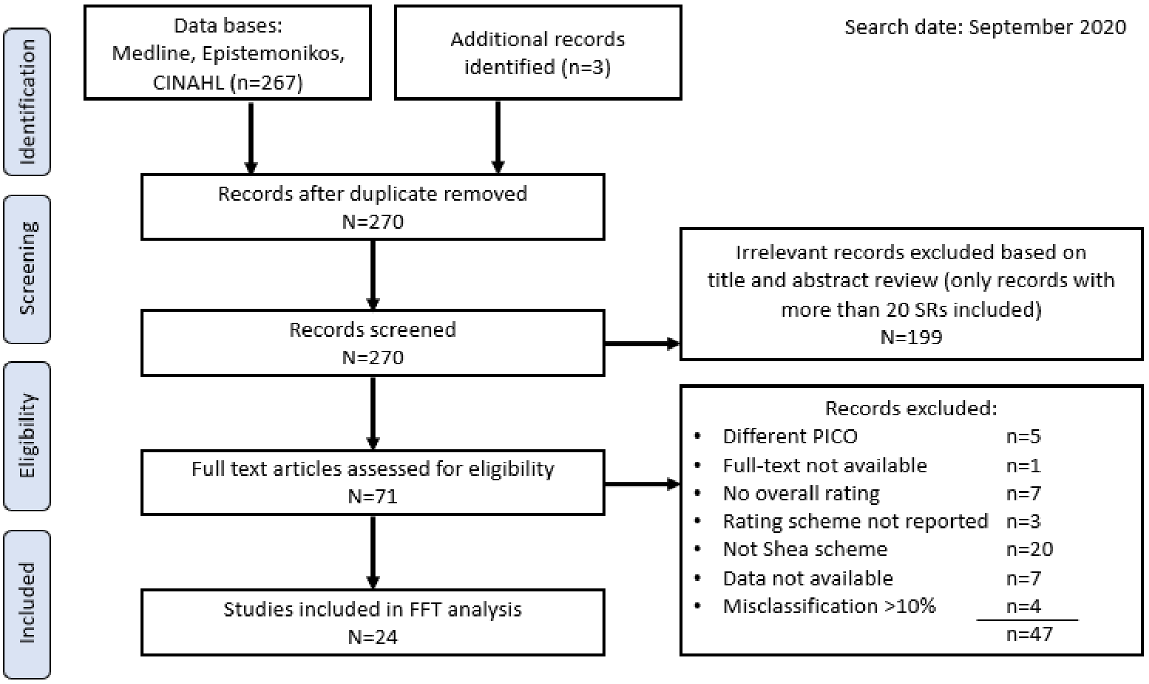

Figure S1: Modified PRISMA flow diagram

### *List of included studies*

Almeida, M. O., Yamato, T. P., Parreira, P., Costa, L. O. P., Kamper, S., & Saragiotto, B. T. (2020). Overall confidence in the results of systematic reviews on exercise therapy for chronic low back pain: a cross-sectional analysis using the Assessing the Methodological Quality of Systematic Reviews (AMSTAR) 2 tool. Braz J Phys Ther, 24(2), 103-117. https://doi.org/10.1016/j.bjpt.2019.04.004

Barbosa, A., Whiting, S., Mendes, R., Whiting, S., Mendes, R., Breda, J., Whiting, S., Simmonds, P., Moreno, R. S., Mendes, R., & Breda, J. (2020). Physical activity and academic achievement: An umbrella review. Int. J. Environ. Res. Public Health, 17(16), 1-29. https://doi.org/10.3390/ijerph17165972

Dang, A., Chidirala, S., Veeranki, P., & Vallish, B. N. (2020). A Critical Overview of Systematic Reviews of Chemotherapy for Advanced and Locally Advanced Pancreatic Cancer using both AMSTAR2 and ROBIS as Quality Assessment Tools. Rev Recent Clin Trials. https://doi.org/10.2174/1574887115666200902111510

Ding, M., Soderberg, L., Jung, J. H., & Dahm, P. (2020). Low Methodological Quality of Systematic Reviews Published in the Urological Literature (2016-2018). Urology, 138, 5-10. https://doi.org/10.1016/j.urology.2020.01.004

Eze, N. D., Mateus, C., & Cravo Oliveira Hashiguchi, T. (2020). Telemedicine in the OECD: An umbrella review of clinical and cost-effectiveness, patient experience and implementation. PLoS One, 15(8), e0237585. https://doi.org/10.1371/journal.pone.0237585

Fuchs, S., Grössmann, N., Eckhardt, H., Busse, R., & Wild, C. (2018). PET/PET-CT Evidenz zum Bedarf und zur Planung in Deutschland und Österreich: Update 2018 (Einschluss, Trans.; Vol. 12). Universitätsverlag der TU Berlin.

Gao, Y., Cai, Y., Yang, K., Liu, M., Shi, S., Chen, J., Sun, Y., Song, F., Zhang, J., & Tian, J. (2020). Methodological and reporting quality in non-Cochrane systematic review updates could be improved: a comparative study. J Clin Epidemiol, 119, 36-46. https://doi.org/10.1016/j.jclinepi.2019.11.012

Hacke, C., & Nunan, D. (2020). Discrepancies in meta-analyses answering the same clinical question were hard to explain: a meta-epidemiological study. J Clin Epidemiol, 119, 47-56. https://doi.org/10.1016/j.jclinepi.2019.11.015

He, W., Li, M., Zuo, L., Wang, M., Jiang, L., Shan, H., Han, X., Yang, K., & Han, X. (2019). Acupuncture for treatment of insomnia: An overview of systematic reviews. Complement Ther Med, 42, 407-416. https://doi.org/10.1016/j.ctim.2018.12.020

He, W., Wang, M., Jiang, L., Li, M., & Han, X. (2019). Cognitive interventions for mild cognitive impairment and dementia: An overview of systematic reviews. Complement Ther Med, 47, 102199. https://doi.org/10.1016/j.ctim.2019.102199

Leclercq, V., Beaudart, C., Ajamieh, S., Tirelli, E., & Bruyère, O. (2020). Methodological quality of meta-analyses indexed in PsycINFO: leads for enhancements: a meta-epidemiological study. BMJ Open, 10(8), e036349. https://doi.org/10.1136/bmjopen-2019-036349

Li, H., Liu, Y., Luo, D., Ma, Y., Zhang, J., Li, M., Yao, L., Shi, X., Liu, X., & Yang, K. (2019). Ginger for health care: An overview of systematic reviews. Complement Ther Med, 45, 114-123. https://doi.org/10.1016/j.ctim.2019.06.002

Lin, S. S., Liu, C. X., Zhang, J. H., Wang, X. L., & Mao, J. Y. (2020). Efficacy and Safety of Oral Chinese Patent Medicine Combined with Conventional Therapy for Heart Failure: An Overview of Systematic Reviews. Evid Based Complement Alternat Med, 2020, 8620186. https://doi.org/10.1155/2020/8620186

Lorenz, R. C., Matthias, K., Pieper, D., Wegewitz, U., Morche, J., Nocon, M., Rissling, O., Schirm, J., Freitag, S., & Jacobs, A. (2020). AMSTAR 2 overall confidence rating: lacking discriminating capacity or requirement of high methodological quality? J Clin Epidemiol, 119, 142-144. https://doi.org/10.1016/j.jclinepi.2019.10.006

Matthias, K., Rissling, O., Pieper, D., Morche, J., Nocon, M., Jacobs, A., Wegewitz, U., Schirm, J., & Lorenz, R. C. (2020). The methodological quality of systematic reviews on the treatment of adult major depression needs improvement according to AMSTAR 2: A cross-sectional study. Heliyon, 6(9), e04776. https://doi.org/10.1016/j.heliyon.2020.e04776

Mohseni, S., Aalaa, M., Atlasi, R., Mohajeri Tehrani, M. R., Sanjari, M., & Amini, M. R. (2019). The effectiveness of negative pressure wound therapy as a novel management of diabetic foot ulcers: an overview of systematic reviews. J Diabetes Metab Disord, 18(2), 625-641. https://doi.org/10.1007/s40200-019-00447-6

Moore, R. A., Fisher, E., Finn, D. P., Finnerup, N. B., Gilron, I., Haroutounian, S., Krane, E., Rice, A. S. C., Rowbotham, M., Wallace, M., & Eccleston, C. (2020). Cannabinoids, cannabis, and cannabis-based medicines for pain management: an overview of systematic reviews. Pain. https://doi.org/10.1097/j.pain.0000000000001941

Nascimento, D. P., Gonzalez, G. Z., Araujo, A. C., Moseley, A. M., Maher, C. G., & Costa, L. O. P. (2020). Eight Out of Every Ten Abstracts of Low Back Pain Systematic Reviews Presented Spin and Inconsistencies With the Full Text: An Analysis of 66 Systematic Reviews. J Orthop Sports Phys Ther, 50(1), 17-23. https://doi.org/10.2519/jospt.2020.8962

Smires, S., Afach, S., Mazaud, C., Phan, C., Doval, I. G., Boyle, R., Dellavalle, R., Williams, H. C., Grindlay, D., Sbidian, E., & Le Cleach, L. (2020). Quality and reporting completeness of systematic reviews and meta-analyses in dermatology. The Journal of investigative dermatology. https://doi.org/10.1016/j.jid.2020.05.109

Smith, A. L., Brown, J., Wyman, J. F., Berry, A., Newman, D. K., & Stapleton, A. E. (2018). Treatment and Prevention of Recurrent Lower Urinary Tract Infections in Women: A Rapid Review with Practice Recommendations. J Urol, 200(6), 1174-1191. https://doi.org/10.1016/j.juro.2018.04.088

Smith, V., Gallagher, L., Carroll, M., Hannon, K., & Begley, C. (2019). Antenatal and intrapartum interventions for reducing caesarean section, promoting vaginal birth, and reducing fear of childbirth: An overview of systematic reviews. PLoS One, 14(10), e0224313. https://doi.org/10.1371/journal.pone.0224313

Snowdon, N., Allan, J., Shakeshaft, A., Rickwood, D., Stockings, E., Boland, V. C., & Courtney, R. J. (2019). Outpatient psychosocial substance use treatments for young people: An overview of reviews. Drug Alcohol Depend, 205, 107582. https://doi.org/10.1016/j.drugalcdep.2019.107582

Zhang, J., Zhang, Y., Huang, X., Lan, K., Hu, L., Chen, Y., & Yu, H. (2020). Different Acupuncture Therapies for Allergic Rhinitis: Overview of Systematic Reviews and Network Meta-Analysis. Evid Based Complement Alternat Med, 2020, 8363027. https://doi.org/10.1155/2020/8363027

Zhou, C., Zhong, X., Song, Y., Shi, J., Wu, Z., Guo, Z., Sun, J., & Wang, Z. (2019). Prognostic Biomarkers for Gastric Cancer: An Umbrella Review of the Evidence. Front Oncol, 9, 1321. https://doi.org/10.3389/fonc.2019.01321

#### *Alternative Short appraisal FFT*

Since the sensitivity in the short appraisal FFT was only 80%, the decision tree with the highest balanced accuracy (92%) is presented here. The tree is only slightly different from the FFT that is presented in the main text of the article. The order of the selected items is the same, but the exit leave in the second level (item 2, pre-defined protocol) is different: exit high at a yes. See also: Figure S1.

The accuracy was 92% with 89 hits, 605 correct rejections, 5 misses and 59 false alarms leading to a sensitivity of 95% and a specificity of 91%. The positive predictive value was 60 % and the negative predictive value was 99%. In this FFT the mcu (mean cues used) was 1.4 and the pci (percentage of cues ignored) was 92%.

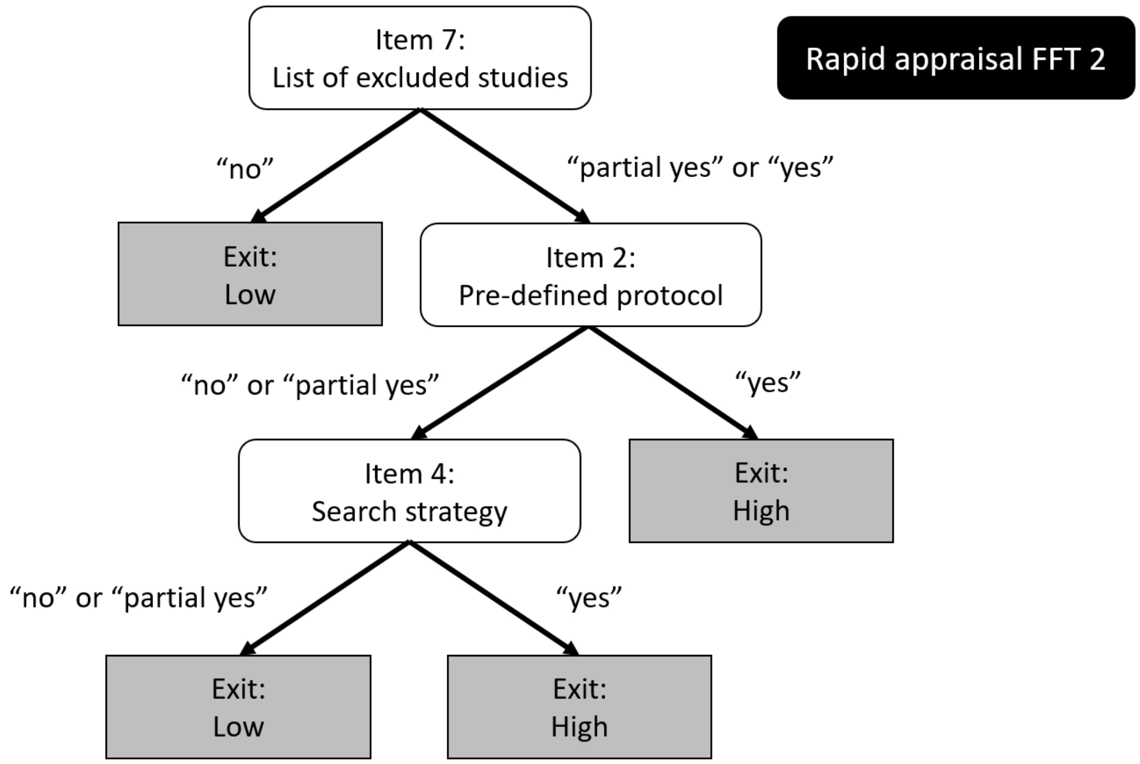

Figure S2: The fast-and-frugal decision tree for short appraisal is shown (alternative tree to the presented tree in the main text). This tree contains three items of the AMSTAR 2 that are ordered the following way: On the first level item 7 (list of excluded studies), on the second level item 2 (pre-defined protocol) and on the final level item 4 (search strategy).

### *Characteristics of the included studies*

*Table S2: Characteristics of the included studies including number of AMSTAR 2 overall confidence ratings.*

| Reference | Type of article | Clinical field | Number of AMSTAR 2 appraisals | High | Mode-rate | Low | Critically low |
|---|---|---|---|---|---|---|---|
| Almeida et al. 2020 | methods study | diseases of the musculoskeletal system | 38 | 3 | 1 | 6 | 28 |
| Barbosa et al. 2020 | overview of systematic reviews | diverse fields | 41 | 2 | 5 | 11 | 23 |
| Dang et al. 2020 | overview of systematic reviews | oncological diseases | 26 | 0 | 0 | 1 | 25 |
| Ding et al. 2020 | methods study | diseases of the genitourinary system | 144 | 6 | 1 | 31 | 106 |
| Eze et al. 2020 | overview of systematic reviews | diverse fields | 98 | 8 | 0 | 29 | 61 |
| Fuchs et al. 2018 | HTA report | diverse fields | 22 | 0 | 2 | 1 | 19 |
| Gao et al. 2020 | methods study | diverse fields | 60 | 0 | 0 | 2 | 58 |
| Hacke et al. 2020 | methods study | diverse fields | 44 | 2 | 15 | 10 | 16 |
| He, Li et al. 2019 | overview of systematic reviews | mental illness | 34 | 2 | 0 | 21 | 11 |
| He, Wang et al. 2019 | overview of systematic reviews | mental illness | 22 | 0 | 0 | 2 | 20 |
| Leclercq et al., 2020 | methods study | mental illness | 207 | 1 | 2 | 8 | 196 |
| Li et al., 2019 | overview of systematic reviews | diverse fields | 27 | 0 | 2 | 5 | 20 |
| Lin et al., 2020 | overview of systematic reviews | diseases of the cardiovascular system | 38 | 0 | 0 | 0 | 38 |
| Lorenz et al., 2020 | methods study | mental illness | 58 | 2 | 6 | 8 | 42 |
| Matthias et al., 2020 | methods study | mental illness | 60 | 4 | 2 | 1 | 53 |
| Mohseni et al., 2019 | overview of systematic reviews | metabolic diseases | 23 | 7 | 0 | 8 | 8 |
| Moore et al., 2020 | overview of systematic reviews | diverse fields | 57 | 2 | 6 | 8 | 41 |

| | | | | | | | |
|---|---|---|---|---|---|---|---|
| Nascimento et al., 2020 | methods study | diseases of the musculoskeletal system | 66 | 5 | 4 | 7 | 50 |
| Smires et al., 2020 | methods study | diverse fields | 140 | 8 | 1 | 5 | 126 |
| Smith et al., 2018 | overview of systematic reviews | diseases of the genitourinary system | 23 | 3 | 0 | 5 | 15 |
| Smith et al., 2019 | overview of systematic reviews | gynaecology | 155 | 66 | 14 | 21 | 54 |
| Snowdon et al., 2019 | overview of systematic reviews | mental illness | 40 | 6 | 1 | 1 | 32 |
| Zhang et al., 2020 | overview of systematic reviews | diseases of the respiratory system | 20 | 0 | 0 | 3 | 17 |
| Zhou et al, 2019 | overview of systematic reviews | oncological diseases | 76[1] | 0 | 0 | 0 | 76 |

[1] In the publication from Zhou et al., 2019 were biomarkers of gastric cancer were investigated. 74 reviews were included in the overview of systematic reviews, but more critical appraisals of AMSTAR2 were included, because in some cases a critical appraisal was conducted per biomarker. In some cases, more biomarkers were included in the overview of systematic reviews.
